# Assessing treatment effect modification due to comorbidity using individual participant data from industry-sponsored drug trials

**DOI:** 10.1101/2023.01.19.23284762

**Authors:** Peter Hanlon, Elaine W Butterly, Anoop SV Shah, Laurie J Hannigan, Jim Lewsey, Frances S Mair, David Kent, Bruce Guthrie, Sarah H Wild, Nicky J Welton, Sofia Dias, David A McAllister

## Abstract

**Background:** People with comorbidities are under-represented in clinical trials. Empirical estimates of treatment effect modification by comorbidity are lacking leading to uncertainty in treatment recommendations. We aimed to produce estimates of treatment effect modification by comorbidity using individual participant data (IPD).

**Methods and Results:** Using 126 industry-sponsored phase 3/4 trials across 23 index conditions, we performed a two-stage IPD meta-analysis to estimate modification of treatment effect by comorbidity. We estimated the effect of comorbidity measured in 3 ways: (i) the number of comorbidities (in addition to the index condition), (ii) presence or absence of the six commonest comorbid diseases for each index condition, and (iii) using continuous markers of underlying conditions (e.g., estimated glomerular function).

Comorbidities were under-represented in trial participants and few had >2 comorbidities. We found no evidence of modification of treatment efficacy by comorbidity, for any of the 3 measures of comorbidity. This was the case for 20 conditions for which the outcome variable was continuous (e.g., change in glycosylated haemoglobin in diabetes) and for three conditions in which the outcomes were discrete events (e.g., number of headaches in migraine). Although all were null, estimates of treatment effect modification were more precise in some cases (e.g., Sodium-glucose co-transporter inhibitors for type 2 diabetes – interaction term for comorbidity count 0.004, 95% CI - 0.01 to 0.02) while for others credible intervals were wide (e.g., corticosteroids for asthma – interaction term -0.22, 95% CI -1.07 to 0.54).

**Conclusion:** For trials included in this analysis, there was no empirical evidence of treatment effect modification by comorbidity. Our findings support the assumption that estimates of treatment efficacy are constant, at least across modest levels of comorbidity.

## Introduction

Multimorbidity, the presence of two or more long-term conditions, is a global clinical and public health priority.^1,2^ The prevalence of multimorbidity is such that most people with a given long term condition also have comorbidities (referring to additional long-term conditions in the context of an index condition). There is uncertainty about how individual long-term conditions should be managed in the presence of comorbidities.^3^ A major driver of this uncertainty is the underrepresentation of people with multimorbidity in randomised controlled trials.^4,5^ Trial populations are typically younger, healthier and have fewer comorbidities than people treated in routine clinical practice. This has led clinical guideline developers to caution against the application of single-disease recommendations for people with multimorbidity.^6^ However, despite the challenges to clinical management posed by this uncertainty, the efficacy of treatments in the context of comorbidity is rarely assessed. It is therefore not clear, for most treatments, whether relative treatment efficacy differs in people with comorbidity.

Assessing individual differences in response to medical treatments is a controversial topic. Differences in treatment efficacy is typically assessed using subgroup analyses. Subgroup analyses in randomised controlled trials (RCTs) seek to assess if treatment efficacy differs by patient characteristics.^7^ Testing of pre-specified subgroup effects is common practice in RCTs of medical therapies.^8,9^ As such, subgroup analyses seek to inform stratified approaches to patient care by identifying groups for whom recommendations may be tailored.^10^ However, trials rarely report subgroup analyses by levels of comorbidity or for specific comorbidities. Furthermore, subgroup analyses are inconsistently executed and reported, as well as suffering a number of well-documented statistical pitfalls,^7,11^ notably that analysis of subgroups risks false positive and false negative findings.^11^ RCTs are generally not powered to detect subgroup effects, and as such the sample size in subgroup analyses is frequently insufficient to detect clinically significant differences in treatment efficacy even if these were to exist.^12^ Conversely, by testing multiple subgroups, the likelihood of chance findings (i.e., false positives) is increased.^7,12^

The limitations of trial-level subgroup analyses can be reduced using meta-analyses. However, when considering whether treatment efficacy varies by comorbidity, traditional study-level meta-analysis of published findings are likely to be inadequate as trials rarely report subgroup effects by comorbidity, and those that do may be subject to publication bias. Individual-participant data meta-analysis has the potential to overcome these problems. We previously demonstrated, using data from >100 industry-sponsored clinical trials, that it was possible to identify comorbidities in most trials and that multimorbidity was common (although under-represented) in trial populations.^4^ Furthermore, in a recent simulation study we demonstrated that combining trials on all comparisons for a given indication in Bayesian hierarchical models has several desirable properties in terms of estimating treatment effect modification by comorbidity.^13^ First, precision is higher compared to single-comparison meta-analyses, increasing the likelihood of detecting small (but clinically relevant) subgroup effects where these are present. Secondly, extreme values are attenuated towards the null (shrinkage), reducing the risk of false positive findings.^13^ Bayesian hierarchical models may therefore be a useful tool to assess treatment efficacy estimates in the context of multimorbidity.

This study aims to assess whether treatment effects are modified in the presence of comorbidity, by using individual participant data (IPD) from 126 trials to assess whether treatment efficacy for 23 index conditions differs by i) age and sex, ii) number of additional long-term conditions (comorbidity count), (iii) the six commonest comorbidities for each index condition, (iv) by continuous biomarkers associated with comorbidity.

## Methods

### Study design

For trials of 23 index conditions we identified comorbid long-term conditions using IPD for each trial. We then summarised these as a comorbidity count (in addition to the index condition) for each participant. Further, we identified the six commonest comorbidities for each index condition across trials and defined a presence/absence variable for each. We estimated differences in treatment efficacy by fitting regression models to IPD for each trial to obtain trial-level estimates of covariate-treatment interaction effects. We fit models for age and sex alone, for a comorbidity count, and for each of the six commonest comorbidities for each index condition. Trial level estimates were then meta-analysed to obtain drug- and index-condition specific estimates of treatment effect modification by comorbidity. This process is summarised in Figure 1 and explained in detail below.

**Figure 1:**
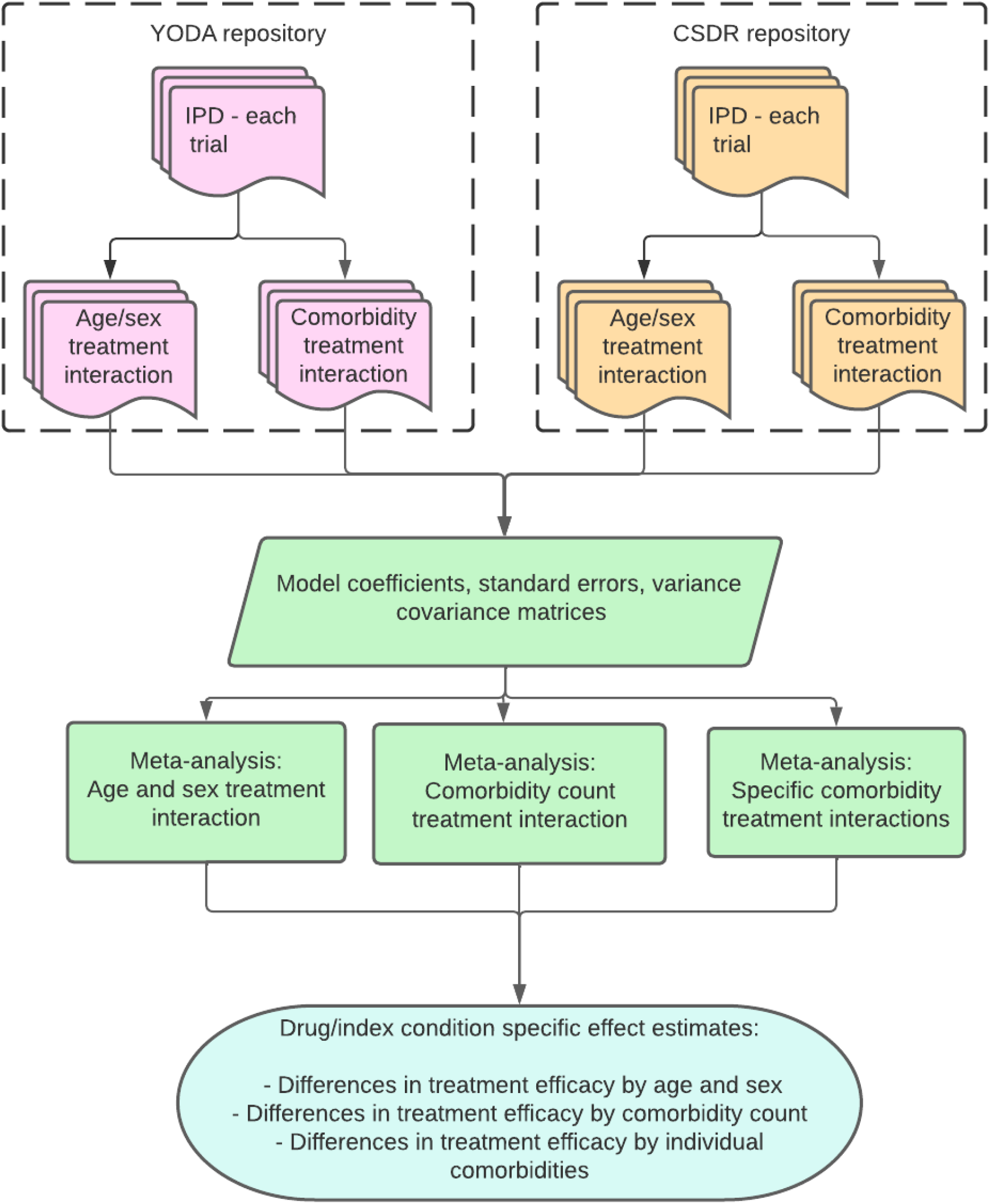
Overview of analysis

All analyses were conducted in R (R Core Team, 2021). Analysis code, metadata (indicating, for example, how treatment arms and outcomes were selected) and data (except trial IPD) are available on the project github repository).

### Data sources

Trials were identified according to a pre-specified protocol.^14^ We focused on trials of pharmacological agents for 23 index conditions (Table 1). Eligibility criteria were industry-sponsored RCTs for one of the index conditions; registered with the United States Clinical trials registry (clinicaltrials.gov) on or after January 1990; phase 2/3, 3, or 4; including ≥ 300 participants, and with eligibility defined using an upper age limit of 60 years or more or no upper age limit. Smaller studies and studies with lower age limits were excluded as they were considered less likely to include sufficient people with comorbidity. From a list of all registered, eligible trials we then identified trials for which IPD were available from one of two repositories: Clinical Study Data Request (CSDR) or the Yale Open Data Access (YODA) repository. The process of trial identification is described in detail elsewhere.^4^

**Table 1:** Index conditions, outcomes and treatment comparisons for included trials

| Index conditions | Outcome | Treatment comparisons | Trials |
| --- | --- | --- | --- |
| Trials with continuous outcomes |  |  |  |
| Ankylosing Spondylitis | BASDAI score | Interleukin inhibitors (L04AC)-IL6 [1],<br>Tumor necrosis factor alpha (TNF-) inhibitors (L04AB) [2] | 3 |
| Asthma | FEV1 | Glucocorticoids (R03BA) [2],<br>Other systemic drugs for obstructive airway diseases (R03DX) [1],<br>Selective beta-2-adrenoreceptor agonists (R03AC) [1] | 4 |
| Benign Prostatic Hypertrophy | IPSS Total Score | Drugs used in erectile dysfunction (G04BE) [5] | 5 |
| Chronic Idiopathic Urticaria | DLQI Score | Other systemic drugs for obstructive airway diseases (R03DX) [3] | 3 |
| Dementia | ADAS score | Anticholinesterases (N06DA) [3] & Thiazolidinediones (A10BG) [3] | 6 |
| Diabetes | HBA1c | Biguanides (A10BA) vs Glucagon-like peptide-1 (GLP-1) analogues (A10BJ) [1],<br>Dipeptidyl peptidase 4 (DPP-4) inhibitors (A10BH) [1],<br>Dipeptidyl peptidase 4 (DPP-4) inhibitors (A10BH) vs Glucagon-like peptide-1 (GLP-1) analogues (A10BJ) [1],<br>Dipeptidyl peptidase 4 (DPP-4) inhibitors (A10BH) vs Sodium-glucose co-transporter 2 (SGLT2) inhibitors (A10BK) [1],<br>Glucagon-like peptide-1 (GLP-1) analogues (A10BJ) [2],<br>Insulins and analogues (A10A) vs Glucagon-like peptide-1 (GLP-1) analogues (A10BJ) [2],<br>Sodium-glucose co-transporter 2 (SGLT2) inhibitors (A10BK) [12],<br>Sulfonylureas (A10BB) vs Dipeptidyl peptidase 4 (DPP-4) inhibitors (A10BH) [1],<br>Sulfonylureas (A10BB) vs Sodium-glucose co-transporter 2 (SGLT2) inhibitors (A10BK) [1] | 22 |
| Erectile dysfunction | IPSS Total Score | Drugs used in erectile dysfunction (G04BE) [1] | 1 |
| Gastro-oesophageal Reflux Disease | Percent heartburn free days | Proton pump inhibitors (A02BC) [2] | 2 |
| Gout | Urate | Preparations inhibiting uric acid production (M04AA) [1] | 1 |
| Hypertension | Systolic blood pressure | ACE inhibitors, plain (C09AA) vs Angiotensin II antagonists, plain (C09CA) [3],<br>Angiotensin II antagonists, plain (C09CA) [1],<br>Thiazides & plain (C03AA) [1] | 5 |
| Inflammatory Bowel Disease | CDAI Score or MAYO score | Interleukin inhibitors (L04AC)-IL12-IL23 [3],<br>Selective immunosuppressants (L04AA) [2].<br>Tumor necrosis factor alpha (TNF-) inhibitors (L04AB) [6] | 11 |
| Inflammatory Arthropathy | ACR numerical | Interleukin inhibitors (L04AC)-IL12-IL23 [1],<br>Interleukin inhibitors (L04AC)-IL6 [4],<br>Tumor necrosis factor alpha (TNF-) inhibitors (L04AB) [8],<br>Tumor necrosis factor alpha (TNF-) inhibitors (L04AB) vs Interleukin inhibitors (L04AC)-IL6 [1] | 14 |
| Osteoporosis | BMD Total Hip | Bisphosphonates (M05BA) [2],<br>Bisphosphonates (M05BA) vs Parathyroid hormones and analogues (H05AA) [1],<br>Parathyroid hormones and analogues (H05AA) [2] | 5 |
| Parkinson Disease | UPDRS Total | Dopamine agonists (N04BC) [4] | 4 |
| Psoriasis | PASI score | Interleukin inhibitors (L04AC)-IL12-IL23 [2],<br>Interleukin inhibitors (L04AC)-IL17A [4] | 6 |
| Pulmonary Disease, Chronic Obstructive | FEV1 | Glucocorticoids (R03BA) [3],<br>Selective beta-2-adrenoreceptor agonists (R03AC) [1],<br>Selective beta-2-adrenoreceptor agonists (R03AC) vs Anticholinergics (R03BB) [2] | 6 |
| Pulmonary Fibrosis | FVC | Other protein kinase inhibitors (L01EX) [2] | 2 |
| Restless Legs Syndrome | RLS Symptom Score Total | Dopamine agonists (N04BC) [3] | 3 |
| Rhinitis, allergic | Total Nasal Symptom Score | fluticasone (R01AD08) [1] | 1 |
| Systemic Lupus Erythematosus | SLE Disease Activity Index | Selective immunosuppressants (L04AA) [2] | 2 |
| Trials with categorical outcomes |  |  |  |
| Migraine | No. headaches | topiramate (N03AX11) [5] | 5 |
| Osteoporosis | Vertebral fracture | Parathyroid hormones and analogues (H05AA) [1] & Bisphosphonates (M05BA) [2] | 3 |
| Thromboembolic | Bleeding [1];<br>DVT or PE [1];<br>DVT or PE and Bleeding [7] | Vitamin K antagonists (B01AA) vs Direct thrombin inhibitors (B01AE) [4],<br>Heparin group (B01AB) [1],<br>Heparin group (B01AB) vs Direct thrombin inhibitors (B01AE) [3],<br>Direct thrombin inhibitors (B01AE) [1] | 9 |

*“Treatment comparisons” indicates the treatment comparisons for each trial based on drug class using the WHO ATC code (For L04AC-Interleukin inhibitors the ATC class was also further split according to the specific interleukin(s). Where there is only a single code the comparator is either placebo or usual care).*

### Quantifying comorbidity

For each participant with a specified index condition in each of the included trials, we identified co-morbidities from a pre-specified list of 21 conditions (cardiovascular disease, chronic pain, arthritis, affective disorders, acid-related disorders, asthma/chronic obstructive pulmonary disease, diabetes mellitus, osteoporosis, thyroid disease, thromboembolic disease, inflammatory conditions, benign prostatic hyperplasia, gout, glaucoma, urinary incontinence, erectile dysfunction, psychotic disorders, epilepsy, migraine, parkinsonism, and dementia).^4^ These comorbidities were based on previous work identifying comorbidities within trial IPD, and were based on assessment of medical history and concomitant medication data. In this previous work, we demonstrated that while for many trials medical history had been redacted, data on concomitant medications were widely available, and could be used to define comorbidities.^4^ This involved combining some conditions into the same definition (e.g., asthma and chronic obstructive pulmonary disease, which could not be differentiated based on medication use alone). These definitions were based on the World Health Organisation Anatomic Therapeutic Classification and are described in our previous publication and available on the project github repository.^4^ Where medical history data was available and coded using the Medical Dictionary for Regulatory Activities (MedDRA) coding system, we also identified the same conditions using MedDRA codes.

#### Comorbidity count

For the primary analysis, we created a comorbidity count for each participant. This was the total number of comorbidities present, not including the index condition. This count was used as a numerical variable in all analyses.

#### Individual comorbidities

For each index condition, we also identified the six most common comorbidities from the full list of 21 possible comorbidities. These individual comorbidities were analysed as binary variables (reflecting the presence of absence of that specific comorbidity).

#### Selected biomarkers/risk factors

In addition to the 21 comorbidities defined using medication and/or medical history, we identified five continuous biomarkers which may indicate comorbidity (e.g. renal impairment, hypertension, anaemia or liver disease) or risk factors (e.g. obesity). These were based on baseline trial measurements: estimated Glomerular Filtration Rate (eGFR, as a marker of renal impairment, taken from trial data where this was available and calculated from creatinine, age, sex, and race using the Modification of Diet in Renal Disease (MDRD) equations if it was not), body mass index (as recorded, or calculated based on height and weight), Fibrosis-4 (FIB-4) Index (as a marker of liver disease calculated from aspartate aminotransferase, alanine transaminase and platelet counts), haemoglobin, and mid-blood pressure (MBP, defined as 0.5 x (systolic blood pressure + diastolic blood pressure)).

### Demographics

Age and sex were extracted from each trial based on the trial recorded values at randomisation.

### Treatment arms

Treatment arm comparisons were pre-specified prior to undertaking the outcome analyses. For multi-arm trials the most extreme arms were selected for comparison (e.g., if different dosages were used, the highest dose was compared to placebo or usual care – e.g., canagliflozin 300mg, rather than 100mg, vs placebo). Where placebo or usual care was included as a trial arm, this was selected as the comparator. Otherwise, we chose the arm with the least recently developed treatment as the comparator arm. This was to give the best chance of identifying effect modification, with the resulting analysis representing an upper limit on the degree of effect modification observed.

### Outcomes

We aimed to identify outcomes common across trials to facilitate meta-analysis. We obtained information from clincialtrials.gov via the Database for Aggregated Analysis of ClinicalTrials.gov (AACT; https://ctti-clinicaltrials.org/citation-policy/) on all outcomes (primary and secondary) for each trial. For each index condition we then identified one or more outcomes which appeared to be common to multiple trials (e.g., Forced expiratory volume in 1 second (FEV1) in chronic obstructive pulmonary disease (COPD) trials, 6-minute walk distance (6MWD) in pulmonary hypertension trials). Within the trial repositories we then reviewed the trial documentation to identify these outcomes for each trial.

### Statistical analyses

In four separate analyses we i) estimated age and sex treatment interactions without including comorbidity, ii) estimated comorbidity-treatment interactions for the comorbidity count, iii) estimated comorbidity-treatment interactions for the six commonest comorbidities for each index condition and iv) examined covariate-treatment interactions for continuous biomarkers. Full descriptions of the modelling are provided in the supplementary appendix and are described briefly below.

#### IPD analysis

For trials where the outcome was a continuous variable, for each trial and analysis the change in each outcome was modelled using linear regression. For analysis (i), the final measure was regressed on the baseline measure, age (per 15-year increment, which was close to the standard deviation for most trials), sex (male versus referent group of females) arm (binary variable treatment/control) and interactions with arm for each covariate. For American College of Rheumatology-N (ACR-N, a measure of improvement in disease activity in rheumatoid arthritis, which is itself a measure of change) we did not include the baseline measure as a covariate. We then repeated this modelling for the remaining analyses (ii-iv) adding comorbidity covariates in addition to age and sex (comorbidity count, specific comorbidities, and continuous biomarkers for analyses ii-iv, respectively). From these models, the model coefficients, standard errors, and variance-covariance matrices were obtained and exported from the YODA and CSDR secure analysis platforms.

For trials where the outcome was a count or a binary variable, we fitted similar models using Poisson regression and logistic regression, respectively.

#### Meta-analysis

For the continuous outcomes, in order to convert the measures onto a similar scale we divided the estimates and standard errors by the minimum clinically important difference (MCID) for that measure. For most outcome measures, higher scores indicate worse outcomes (e.g., Bath Ankylosing Spondylitis Disease Activity Index (BASDI)). Where this was not the case (e.g., FEV1) we multiplied the values by minus one so that the direction of effect was the same for all trials. For the variance co-variance-matrix we divided each element by the MCID-squared. The MCID was selected using the published literature by hand-searching papers in the Core Outcome Measures in Effectiveness Trials (COMET) database for relevant conditions.^15^ This search was supplemented by simple internet searches (Google searches using the full and abbreviated names for each outcome and MCID, MID, “minimum clinically important difference” or “minimum important difference”). Where no published MCID recommendations could be found, we used the MCID defined in the power calculations in the trial protocols. At this stage, for each index condition, we restricted the analysis to the single most common outcome across trials. In two index condition (Ankylosing spondylitis and hypertension), two outcomes were equally common; BASDAI and Bath Ankylosing Spondylitis Functional Index (BASFI) and diastolic blood pressure and systolic blood pressure, we arbitrarily chose BASDAI in the former case and chose systolic blood pressure in the latter as it is more prominent in clinical decision making.

For each drug class the model outputs were then meta-analysed. We used random-effects meta-analyses where 5 or more trials were included within the same drug class, and fixed effects where there were fewer than five trials. We used Bayesian models since this allowed us to simultaneously model multiple coefficients (e.g., age-treatment and sex-treatment interactions). The Bayesian models were fit using the *brms* package.(29) Samples from the posterior distribution were obtained and summarised as the mean and 95% credibility intervals (CI).

In case other researchers wish to use the results of our models of treatment-covariate interactions to inform subsequent analyses as informative priors, we obtained summaries of the posterior predictions. We did so only for analysis (ii) for continuous outcomes. In order to provide a more general set of priors, we also predicted the comorbidity count-treatment interaction for treatment comparisons/conditions not included in our model by obtaining samples from the posteriors. We then summarised these samples by fitting a Student’s t-distribution. As with the main analysis, these models were fitted using the *brms* package (see Supplementary appendix for additional details).

## Results

### Trial characteristics

Trial baseline characteristics have been reported previously.^4^ For trials with continuous outcomes there were 20 index conditions and 47 treatment comparisons across a total of 109 trials. For 9 index conditions there was only one treatment comparison across all trials. Diabetes, which was the condition for which there were the most trials (22), had the largest number of treatment comparisons (9) (Table 1). Within each model, all trials had a single common outcome except inflammatory bowel disease, where the ulcerative colitis trials used the MAYO score and Crohn’s disease trials used the Crohn’s Disease Activity Index score. For trials with categorical outcomes there were 3 index conditions (migraine, osteoporosis and thromboembolism) and 11 treatment comparisons across a total of 17 trials, with three index conditions migraine, osteoporosis and thromboembolism. For the latter indication there were three more specific categories of indication - primary prevention (5 trials), secondary prevention (2 trials) and treatment (2 trials).

### Continuous outcomes – age and sex treatment interactions

For all conditions with continuous outcomes, interaction terms for age- and sex-treatment interactions are shown in Table 2. For most drug classes, interaction terms for age included the null, indicating no statistically significant associations consistent with modification of treatment efficacy by age. However, in the diabetes trials, there appeared to be an attenuation in the treatment effect with increasing age for three drug classes (0.07 (95% CI 0.00 to 0.13) for Sulfonylureas vs SGLT2 inhibitors, 0.09 (95% CI 0.01 to 0.17) for DPP-4 inhibitors vs SGLT2 inhibitors and 0.07 (95% CI 0.04 to 0.11) for SGLT2 inhibitors versus placebo). Taking SGLT2 inhibitors versus placebo as an example, this can be read as follows – ‘the lowering effect on HbA1c of SGLT2 inhibitors versus placebo is 0.28 (95% CI 0.16 to 0.44) mmol/mol smaller (since the MCID for HbA1c is 4 mmol/mol) per 15-year increment in age, for age 50 years versus age 80 years this corresponds to the effect being 0.56 (95% CI 0.32 to 0.88) mmol/mol smaller.

**Table 2:**
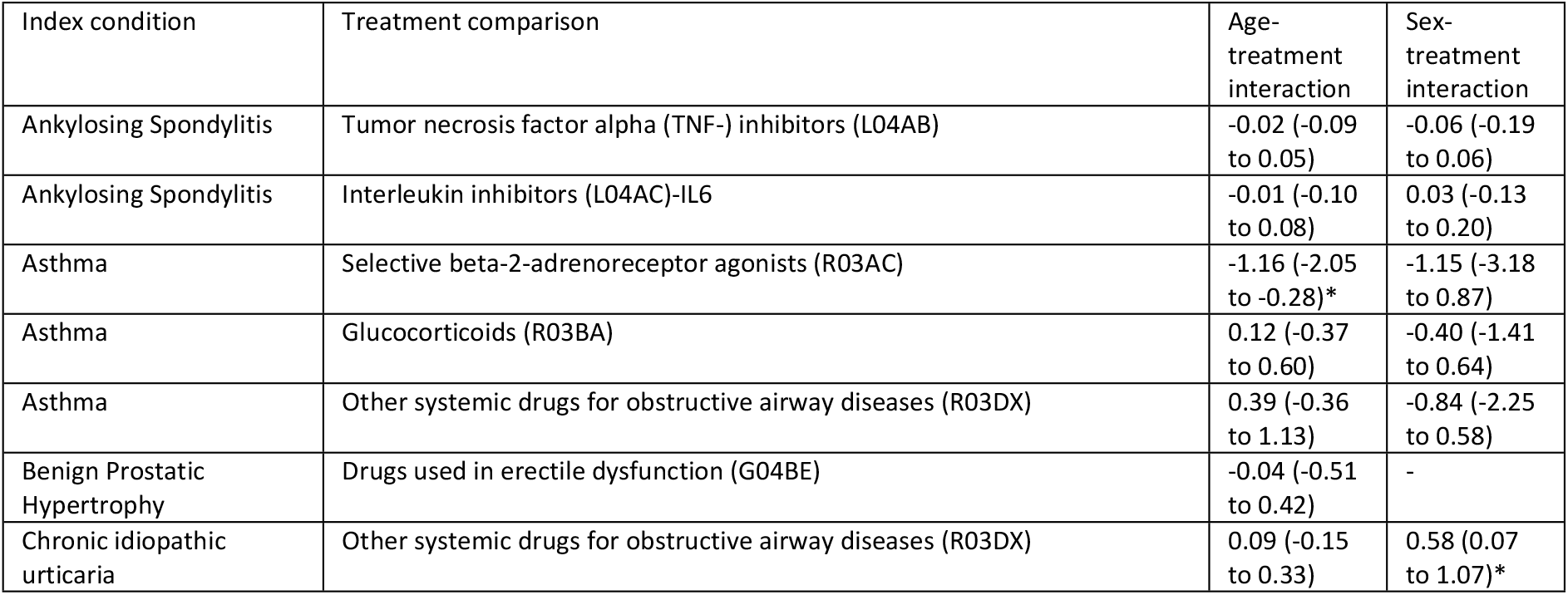
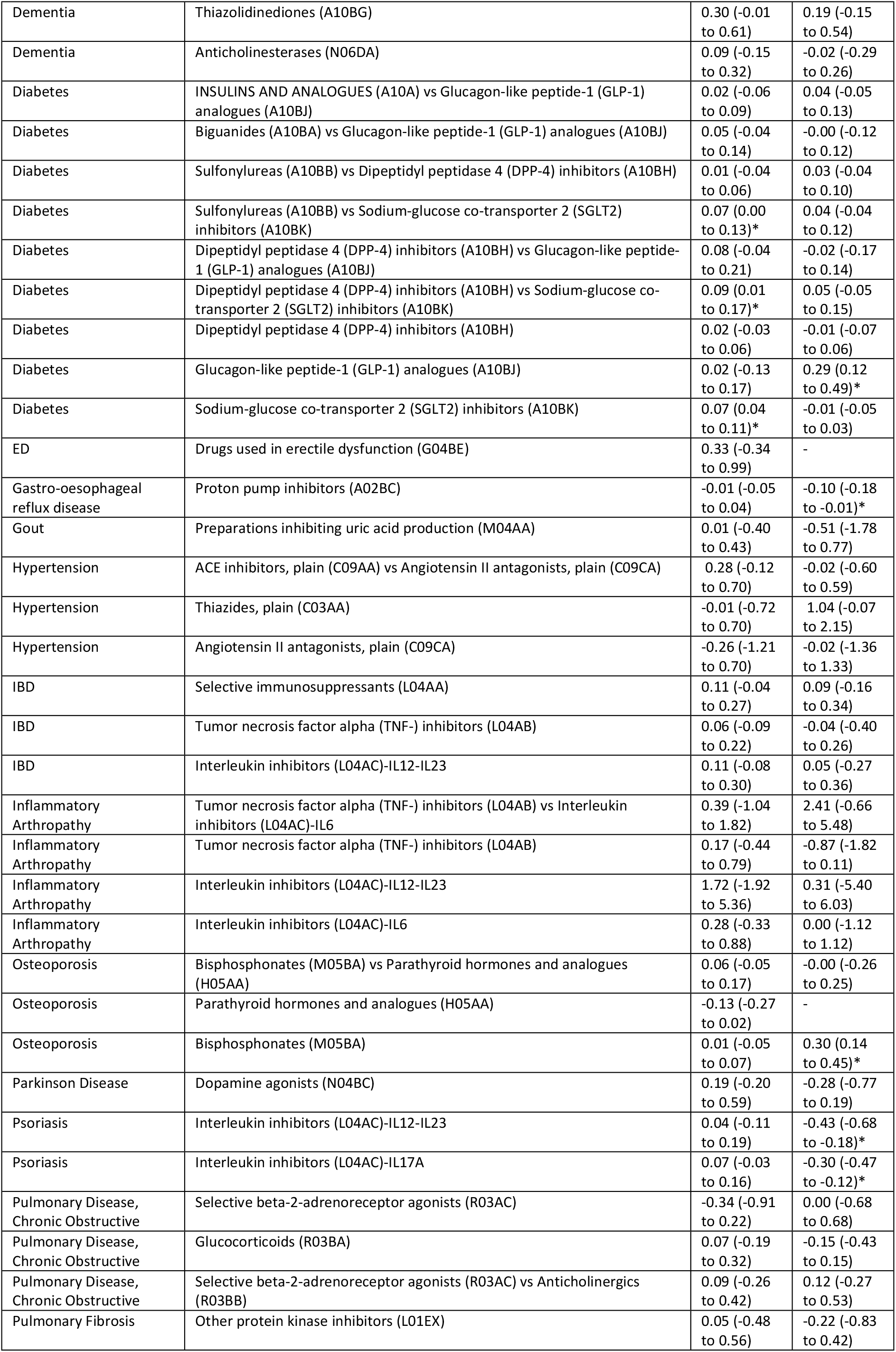
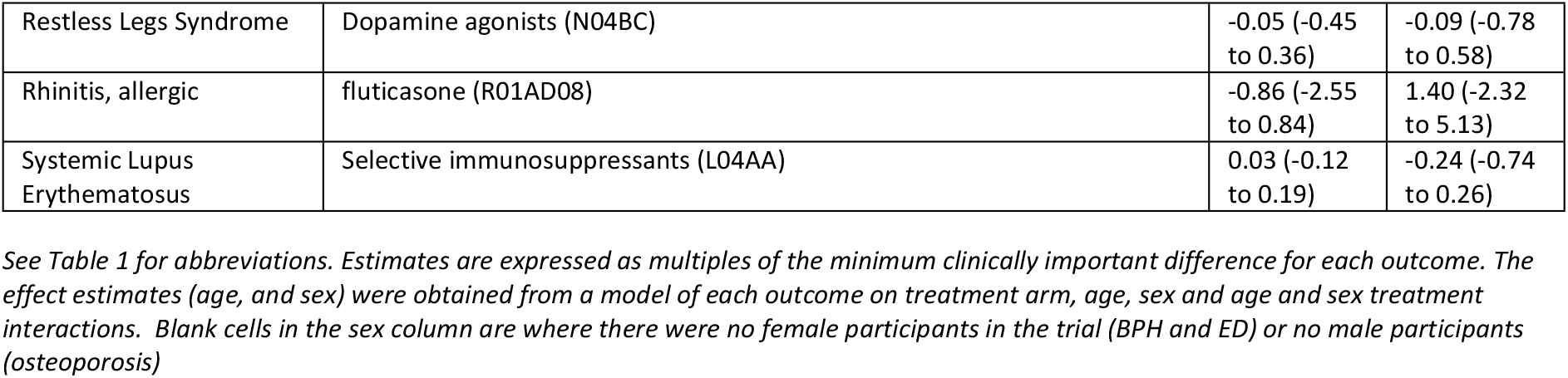
Covariate-treatment interactions (expressed as multiples of minimal clinically important difference) by age and sex for continuous outcomes. Point estimates and 95% credible intervals

| Index condition | Treatment comparison | Age-treatment interaction | Sex-treatment interaction |
| --- | --- | --- | --- |
| Ankylosing Spondylitis | Tumor necrosis factor alpha (TNF-) inhibitors (L04AB) | -0.02 (-0.09 to 0.05) | -0.06 (-0.19 to 0.06) |
| Ankylosing Spondylitis | Interleukin inhibitors (L04AC)-IL6 | -0.01 (-0.10 to 0.08) | 0.03 (-0.13 to 0.20) |
| Asthma | Selective beta-2-adrenoreceptor agonists (R03AC) | -1.16 (-2.05 to -0.28)* | -1.15 (-3.18 to 0.87) |
| Asthma | Glucocorticoids (R03BA) | 0.12 (-0.37 to 0.60) | -0.40 (-1.41 to 0.64) |
| Asthma | Other systemic drugs for obstructive airway diseases (R03DX) | 0.39 (-0.36 to 1.13) | -0.84 (-2.25 to 0.58) |
| Benign Prostatic Hypertrophy | Drugs used in erectile dysfunction (G04BE) | -0.04 (-0.51 to 0.42) | - |
| Chronic idiopathic urticaria | Other systemic drugs for obstructive airway diseases (R03DX) | 0.09 (-0.15 to 0.33) | 0.58 (0.07 to 1.07)* |
| Dementia | Thiazolidinediones (A10BG) | 0.30 (-0.01 to 0.61) | 0.19 (-0.15 to 0.54) |
| Dementia | Anticholinesterases (N06DA) | 0.09 (-0.15 to 0.32) | -0.02 (-0.29 to 0.26) |
| Diabetes | INSULINS AND ANALOGUES (A10A) vs Glucagon-like peptide-1 (GLP-1) analogues (A10BJ) | 0.02 (-0.06 to 0.09) | 0.04 (-0.05 to 0.13) |
| Diabetes | Biguanides (A10BA) vs Glucagon-like peptide-1 (GLP-1) analogues (A10BJ) | 0.05 (-0.04 to 0.14) | -0.00 (-0.12 to 0.12) |
| Diabetes | Sulfonylureas (A10BB) vs Dipeptidyl peptidase 4 (DPP-4) inhibitors (A10BH) | 0.01 (-0.04 to 0.06) | 0.03 (-0.04 to 0.10) |
| Diabetes | Sulfonylureas (A10BB) vs Sodium-glucose co-transporter 2 (SGLT2) inhibitors (A10BK) | 0.07 (0.00 to 0.13)* | 0.04 (-0.04 to 0.12) |
| Diabetes | Dipeptidyl peptidase 4 (DPP-4) inhibitors (A10BH) vs Glucagon-like peptide-1 (GLP-1) analogues (A10BJ) | 0.08 (-0.04 to 0.21) | -0.02 (-0.17 to 0.14) |
| Diabetes | Dipeptidyl peptidase 4 (DPP-4) inhibitors (A10BH) vs Sodium-glucose co-transporter 2 (SGLT2) inhibitors (A10BK) | 0.09 (0.01 to 0.17)* | 0.05 (-0.05 to 0.15) |
| Diabetes | Dipeptidyl peptidase 4 (DPP-4) inhibitors (A10BH) | 0.02 (-0.03 to 0.06) | -0.01 (-0.07 to 0.06) |
| Diabetes | Glucagon-like peptide-1 (GLP-1) analogues (A10BJ) | 0.02 (-0.13 to 0.17) | 0.29 (0.12 to 0.49)* |
| Diabetes | Sodium-glucose co-transporter 2 (SGLT2) inhibitors (A10BK) | 0.07 (0.04 to 0.11)* | -0.01 (-0.05 to 0.03) |
| ED | Drugs used in erectile dysfunction (G04BE) | 0.33 (-0.34 to 0.99) | - |
| Gastro-oesophageal reflux disease | Proton pump inhibitors (A02BC) | -0.01 (-0.05 to 0.04) | -0.10 (-0.18 to -0.01)* |
| Gout | Preparations inhibiting uric acid production (M04AA) | 0.01 (-0.40 to 0.43) | -0.51 (-1.78 to 0.77) |
| Hypertension | ACE inhibitors, plain (C09AA) vs Angiotensin II antagonists, plain (C09CA) | 0.28 (-0.12 to 0.70) | -0.02 (-0.60 to 0.59) |
| Hypertension | Thiazides, plain (C03AA) | -0.01 (-0.72 to 0.70) | 1.04 (-0.07 to 2.15) |
| Hypertension | Angiotensin II antagonists, plain (C09CA) | -0.26 (-1.21 to 0.70) | -0.02 (-1.36 to 1.33) |
| IBD | Selective immunosuppressants (L04AA) | 0.11 (-0.04 to 0.27) | 0.09 (-0.16 to 0.34) |
| IBD | Tumor necrosis factor alpha (TNF-) inhibitors (L04AB) | 0.06 (-0.09 to 0.22) | -0.04 (-0.40 to 0.26) |
| IBD | Interleukin inhibitors (L04AC)-IL12-IL23 | 0.11 (-0.08 to 0.30) | 0.05 (-0.27 to 0.36) |
| Inflammatory Arthropathy | Tumor necrosis factor alpha (TNF-) inhibitors (L04AB) vs Interleukin inhibitors (L04AC)-IL6 | 0.39 (-1.04 to 1.82) | 2.41 (-0.66 to 5.48) |
| Inflammatory Arthropathy | Tumor necrosis factor alpha (TNF-) inhibitors (L04AB) | 0.17 (-0.44 to 0.79) | -0.87 (-1.82 to 0.11) |
| Inflammatory Arthropathy | Interleukin inhibitors (L04AC)-IL12-IL23 | 1.72 (-1.92 to 5.36) | 0.31 (-5.40 to 6.03) |
| Inflammatory Arthropathy | Interleukin inhibitors (L04AC)-IL6 | 0.28 (-0.33 to 0.88) | 0.00 (-1.12 to 1.12) |
| Osteoporosis | Bisphosphonates (M05BA) vs Parathyroid hormones and analogues (H05AA) | 0.06 (-0.05 to 0.17) | -0.00 (-0.26 to 0.25) |
| Osteoporosis | Parathyroid hormones and analogues (H05AA) | -0.13 (-0.27 to 0.02) | - |
| Osteoporosis | Bisphosphonates (M05BA) | 0.01 (-0.05 to 0.07) | 0.30 (0.14 to 0.45)* |
| Parkinson Disease | Dopamine agonists (N04BC) | 0.19 (-0.20 to 0.59) | -0.28 (-0.77 to 0.19) |
| Psoriasis | Interleukin inhibitors (L04AC)-IL12-IL23 | 0.04 (-0.11 to 0.19) | -0.43 (-0.68 to -0.18)* |
| Psoriasis | Interleukin inhibitors (L04AC)-IL17A | 0.07 (-0.03 to 0.16) | -0.30 (-0.47 to -0.12)* |
| Pulmonary Disease, Chronic Obstructive | Selective beta-2-adrenoreceptor agonists (R03AC) | -0.34 (-0.91 to 0.22) | 0.00 (-0.68 to 0.68) |
| Pulmonary Disease, Chronic Obstructive | Glucocorticoids (R03BA) | 0.07 (-0.19 to 0.32) | -0.15 (-0.43 to 0.15) |
| Pulmonary Disease, Chronic Obstructive | Selective beta-2-adrenoreceptor agonists (R03AC) vs Anticholinergics (R03BB) | 0.09 (-0.26 to 0.42) | 0.12 (-0.27 to 0.53) |
| Pulmonary Fibrosis | Other protein kinase inhibitors (L01EX) | 0.05 (-0.48 to 0.56) | -0.22 (-0.83 to 0.42) |
| Restless Legs Syndrome | Dopamine agonists (N04BC) | -0.05 (-0.45 to 0.36) | -0.09 (-0.78 to 0.58) |
| Rhinitis, allergic | fluticasone (R01AD08) | -0.86 (-2.55 to 0.84) | 1.40 (-2.32 to 5.13) |
| Systemic Lupus Erythematosus | Selective immunosuppressants (L04AA) | 0.03 (-0.12 to 0.19) | -0.24 (-0.74 to 0.26) |

### Continuous outcomes – Comorbidity treatment interactions

For each drug class, Figures 2a-2c show the main treatment effect (black points, expressed as change in minimally clinically important difference) and the estimate for the comorbidity-treatment based on a comorbidity count. Meta-analyses for each drug class are shown in Figures 2 and, for classes where only one trial was analysed, trial-level estimates are shown in Figure 2c. Comorbidity count was not associated with any attenuation or strengthening in treatment efficacy; in all cases the 95% credible intervals included the null. When examining comorbidity-treatment interactions for the six most common comorbidities within each index condition, 95% credible intervals included the null for all estimates (supplementary table 1). Similarly, when assessing modification of treatment efficacy by continuous biomarkers all estimates included the null (supplementary table 2).

**Figure 2:**
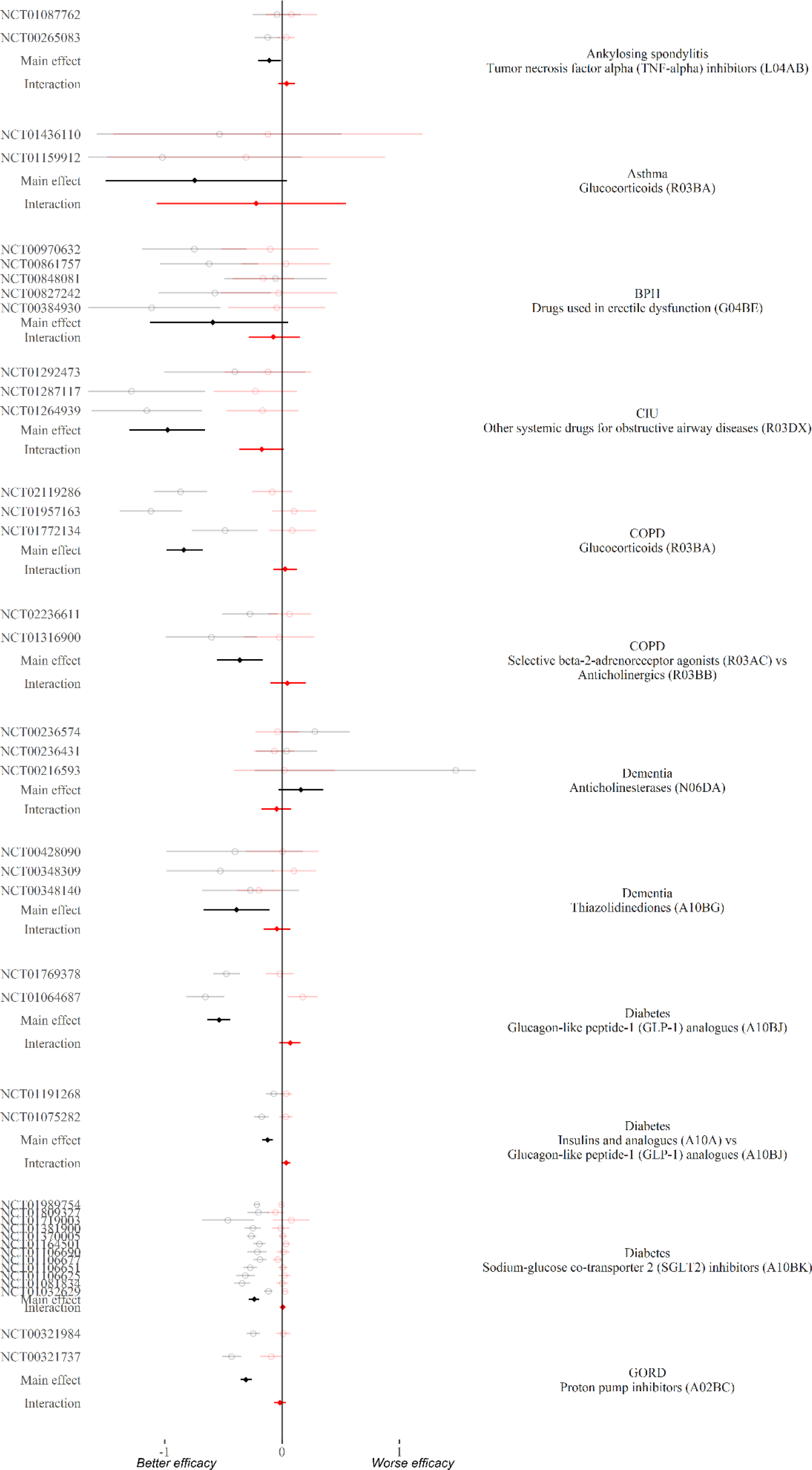
Treatment effect (grey = study level, black = meta-analysis) and comorbidity-treatment interaction (red) based on comorbidity count. Meta-analysis of drug classes.

**Figure 2 :**
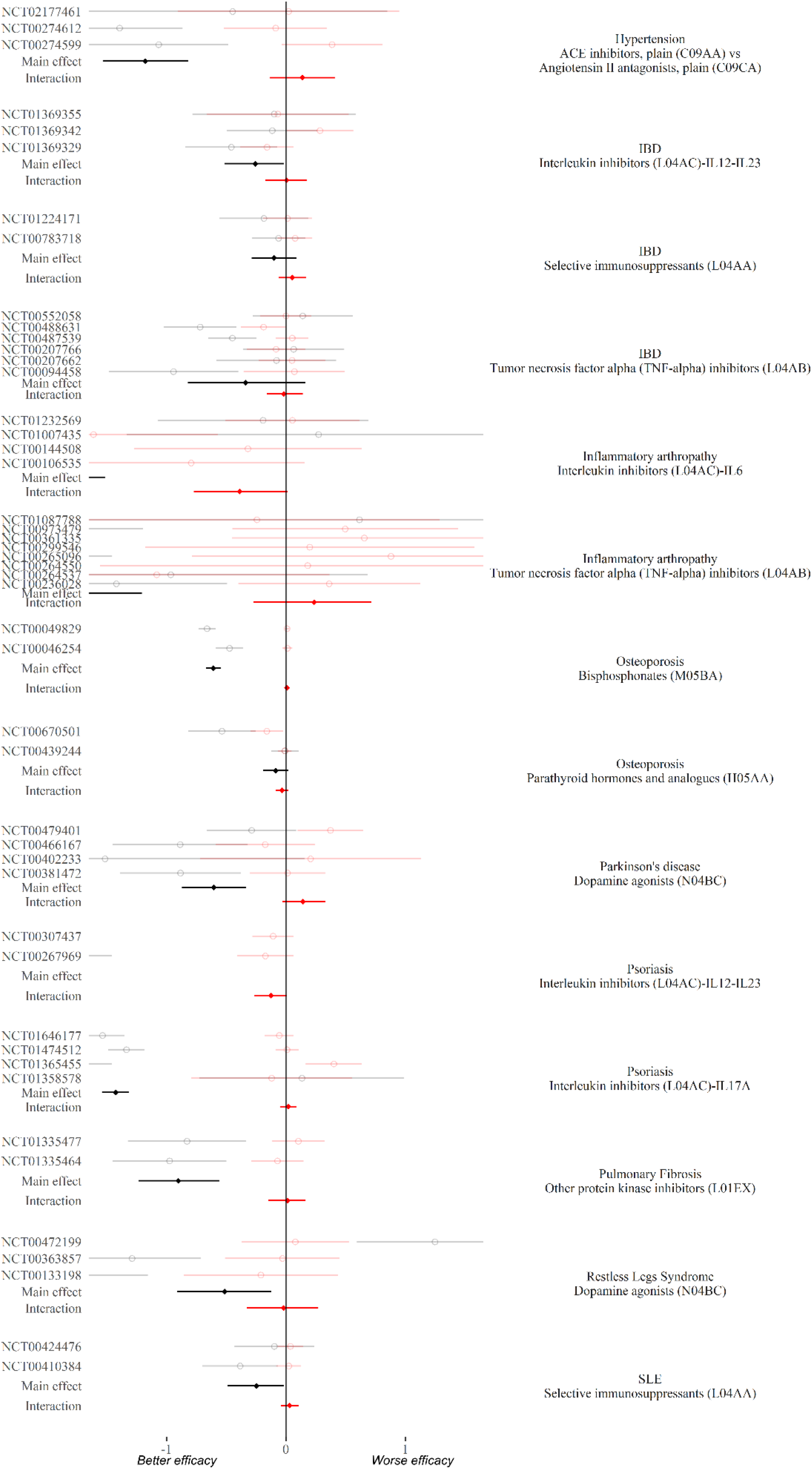
Treatment effect (black) and comorbidity-treatment interaction (red) based on comorbidity count. Meta-analysis of drug classes.

**Figure 2c:**
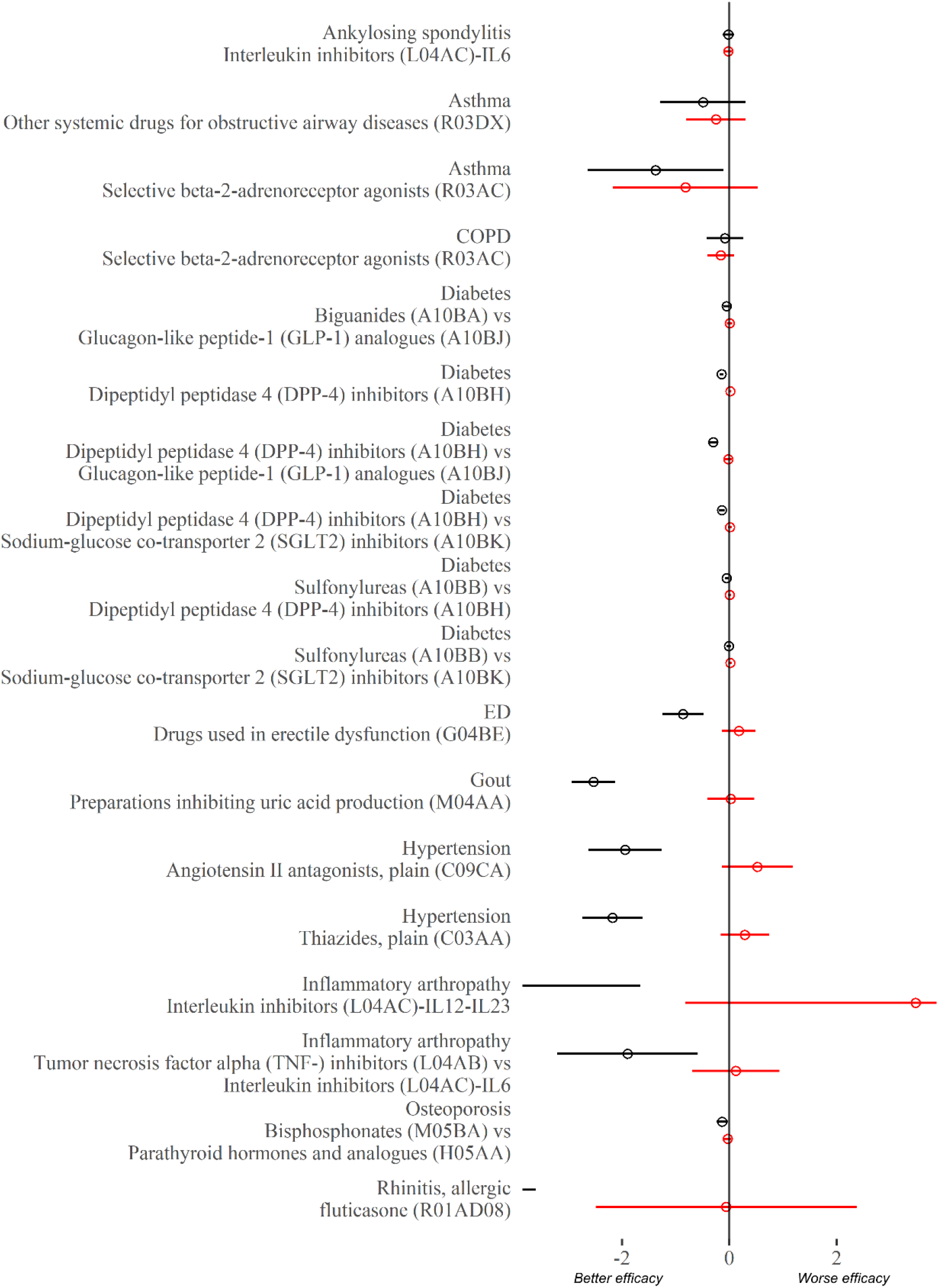
Treatment effect (black) and comorbidity-treatment interaction (red) based on comorbidity count. Single trial estimates.

### Informative priors for subsequent analyses including different index condition/treatment comparisons

On predicting treatment effect modification by comorbidity count for a notional unobserved condition and notional unobserved treatment comparison, the samples from the posterior were approximately t-distributed (central estimate = 0.01, dispersion = 0.01, degrees of freedom = 3.24).

### Categorical outcomes - morbidity-count treatment interactions

For the three index conditions with categorical outcomes (Table 1), there was no evidence of any comorbidity count treatment interactions. These findings are summarised in Table 3.

**Table 3:** Comorbidity-treatment interactions for binary and count outcomes. Point estimates and 95 % credible intervals

| Outcome | Condition | Intervention | Comparator | Trials | Comorbidity count treatment interaction |
| --- | --- | --- | --- | --- | --- |
| Number of headaches | Migraine | topiramate | placebo (N03AX11) | 5 | 1.03 (0.85-1.22) |
| Vertebral fracture | Osteoporosis | teriparatide | placebo (H05AA02) | 1 | 0.87 (0.58-1.30) |
| Vertebral fracture | Osteoporosis | zoledronic acid | placebo (M05BA08) | 2 | 0.96 (0.79-1.17) |
| Bleeding | Primary prevention | dabigatran (B01AE07) | LMWH (B01AB) | 2 | 1.06 (1.00-1.12) |
| DVT/DVT or PE | Primary prevention | dabigatran (B01AE07) | LMWH (B01AB) | 3 | 1.07 (0.89-1.26) |
| Bleeding | Primary prevention | dabigatran (B01AE07) | LMWH (B01AA03) | 1 | 1.00 (0.96-1.04) |
| DVT/DVT or PE | Secondary prevention | dabigatran | placebo (B01AE07) | 1 | 0.82 (0.64-1.05) |
| Bleeding | Secondary prevention | dabigatran | placebo (B01AE07) | 1 | 0.96 (0.60-1.54) |
| DVT/DVT or PE | Secondary prevention | dabigatran (B01AE07) | warfarin (B01AA03) | 1 | 1.06 (0.94-1.19) |
| Bleeding | Secondary prevention | dabigatran (B01AE07) | warfarin (B01AA03) | 1 | 1.05 (0.64-1.70) |
| DVT/DVT or PE | Acute treatment | dabigatran (B01AE07) | warfarin (B01AA03) | 2 | 1.00 (0.90-1.10) |
| Bleeding | Acute treatment | dabigatran (B01AE07) | warfarin (B01AA03) | 2 | 0.91 (0.75-1.10) |

*The interaction estimates represent the ratio per one-unit increase in comorbidity count; effect measures estimates above one indicate worse outcomes in the intervention compared to the comparison arm. For number of migraines the effect estimates are on the rate ratio scale, for the remainder the effect estimates are on the odds ratio scale.*

## Discussion

### Statement of principal findings

In an IPD meta-analysis of 109 trials we examined whether the efficacy of drug treatments differed by comorbidity. For 20 index conditions where the outcome variable was continuous (e.g., glycosylated haemoglobin in diabetes trials), efficacy did not differ by the total number of comorbidities or by the presence or absence of specific comorbidities. Similarly, for 3 conditions (17 trials) examining outcomes which were discrete events (e.g., thromboembolism, bleeding, headaches, and fractures) there was no evidence of treatment effect modification by comorbidity count or by specific comorbidities.

### Strengths and weaknesses of the study

This is the first IPD clinical trial meta-analysis, as far as we are aware, to examine whether treatment efficacy differs by comorbidity. Nonetheless, there are several important limitations. First, while for some index conditions (e.g., diabetes) there were many trials, for others there were few trials and so relatively few participants, limiting the precision with which covariate-treatment interactions could be estimated.

Second, most trials were phase 3 trials focussed on efficacy outcomes (e.g. change in a disease marker such as blood pressure or glycosylated haemoglobin) rather than pragmatic trials focussed on harder outcomes (such as the incidence of specific adverse health outcomes). The findings for the smaller number of trials (17 in total) where we did have harder outcomes (headaches, bleeding, thromboembolism, fracture) were similar to the findings for the remaining trials; there was no evidence of treatment effect modification by comorbidity count on the conventional scale (additive for continuous outcomes and relative for non-continuous outcomes). Nonetheless, the small number of trials and indications where hard outcomes were studied mean that caution is needed in extrapolating our findings to trials or meta-analyses focussing on such outcomes.

Third, while this analysis assesses treatment efficacy we did not assess whether comorbidities lead to variation in adverse effects of treatment. An appreciation of both benefits and harms is required in order to inform judgements about the net benefits of treatment in the context of comorbidity.

Finally, while comorbidity was present in all the included trials, they remain under-representative in terms of the extent comorbidity.^4,16-18^ Specifically, there were few people in the included trials with high comorbidity counts (e.g., 4 or more conditions). This highly multimorbid population is not uncommon in routine clinical practice^19^ and presents considerable challenges for clinical decision making.^3^ Their exclusion from these trials means that our findings cannot be assumed to be directly transferable to patient groups with the highest degree of multimorbidity, for whom uncertainties over the net benefit of treatments are often greatest.^20,21^

### Comparison with other studies

Several previous studies have reported findings on treatment effect modification in IPD meta-analyses and meta-analyses of reported subgroup effects. However, these have largely been confined to major cardiovascular disease trials (e.g., for showing similar efficacy of statin in people with and without diabetes,^22^ or showing questionable net benefit of aspirin in primary prevention^23^) or to concordant conditions defined as those closely related to the index condition or target event for the trial (such as hypertension in stroke trials^24^). These studies have not considered the impact of comorbidity more broadly or of discordant comorbidities not related to the index condition of the trial. This represents an important omission, because there are a number of mechanisms by which the presence of discordant conditions might plausibly modify treatment efficacy (positively or negatively) including increased diagnostic misclassification, altered pharmacokinetics or pharmacodynamics (e.g., altered drug excretion in people with mild renal impairment, or increased benefits of antiplatelet drugs in the presence of co-existent inflammatory conditions) and altered treatment-related behaviours (e.g., better or worse treatment adherence due to existing treatment regimens). Our study adds to this sparse literature showing that, on average, relative treatment effects are similar across different populations within trials (at modest comorbidity counts of 3 or fewer). This supports the standard assumption that treatment effects are similar when generalising from trial to non-trial eligible populations, at least for populations with limited prevalence of comorbidity such as in these trials.

Although we found that relative treatment efficacy did not differ by comorbidity count, net overall treatment benefits may nonetheless differ in people with differing degrees of comorbidity. This is because differences in the baseline risk (e.g., the absolute risk of the outcome that the treatment is intended to prevent), differences in susceptibility to treatment-related adverse events, as well as differences in competing risks (e.g., absolute risk of mortality from other causes) may all lead to differences in the net overall benefit of treatment.^25^ Therefore, where comorbidity alters the (baseline) natural history of diseases, the likelihood of adverse treatment effects (e.g., comorbid renal impairment) or life expectancy (e.g., via discordant comorbidities associated with mortality), the effects of treatment must differ even assuming that there is no difference in efficacy. For example, while there is strong evidence that the benefits of dual antiplatelet therapy (DAPT) following myocardial infarction (versus a single antiplatelet) outweigh the risks,^26,27^ this may not be true for patients with co-existing COPD. Cardiovascular mortality is commoner in COPD than the general population, favouring DAPT.^28^ However, non-cardiovascular mortality is also higher,^29^ favouring single-antiplatelet therapy because of competing risks. Intensive control of blood glucose and other risk factors in diabetes^30,31^ and anticoagulant use in atrial fibrillation,^32^ provide similar examples where the net overall treatment benefits are uncertain for people with comorbidity.

### Implications

In order to estimate net overall treatment benefits, clinical guidelines and health technology assessments routinely use evidence synthesis.^33^ Such approaches combine (i) estimates of relative treatment efficacy with (ii) ‘natural’ history (standard comparator rates) to calculate absolute effectiveness, commonly expressed as the absolute risk reduction (ARR) or number needed to treat.^34^ However, hitherto evidence synthesis has rarely been used to estimate net overall treatment effects for people with multimorbidity. This may partly be due to uncertainty as to whether and how efficacy estimates differ in people with and without comorbidities. Since estimating the natural history rates of target and adverse events for people with multimorbidity is relatively straightforward using routine healthcare data (since such data are sufficiently large and rich in people with multimorbidity to produce such estimates), and within the limitations outlined above, our findings support the standard assumption of estimates of treatment efficacy being constant (at least at the modest levels observed within trial populations).

To support such evidence syntheses, we have provided a set of informative priors which can be used to propagate, into the final treatment effectiveness estimates, the additional uncertainty arising from applying estimates from clinical trials to populations rich in multimorbidity. We summarised the variation in treatment effects by comorbidity count as a set of Student’s t-distributions. These distributions can be used to inform modelling studies (e.g., health technology assessments) designed to extrapolate treatment effect estimates from trial populations to routine clinical practice where multimorbidity is more common. This has the potential to better inform regulatory bodies and guideline developers as they seek to make treatment recommendations for people with multimorbidity.

Our findings also have relevance for analyses of comorbidity sub-group findings in both single clinical trials and as part of meta-analyses. The lack of information for estimating sub-group effects in clinical trials and dangers of falsely claiming spurious sub-group effects is well established and a range of approaches have been advocated for dealing with this problem. These include limiting the number of sub-groups and performing corrections for multiple testing (e.g., the Bonferroni technique used in frequentist analysis), the analysis of treatment effect modification according to participant’s prognostic risk scores at baseline (which reduce the dimensionality of the problem and prioritises characteristics known to predict differences in the rates of target events)^35^ and, in a Bayesian context the use of subject-matter expert knowledge (via prior elicitation). The prior distributions derived from our modelling for the comorbidity-treatment interactions can help inform such prior-elicitation exercises. Another technique used in Bayesian subgroup analyses is to use off-the-shelf conservative priors designed to avoid over fitting;^36^ our findings will help provide reassurance that such priors are unlikely to be overly conservative for modelling comorbidity-treatment interactions.

Finally, our results have relevance for reporting of clinical trial results. Both comorbidity and frailty can be measured using data already collected from clinical trials and – as we show – it is feasible to estimate comorbidity-treatment interactions using such measures. In our project this required access to IPD a process which is expensive (in terms of analysis time) and complex (requiring formal contractual agreements). The PATH statement advocated that clinical trials should report treatment effect modification by baseline prognostic risk score.^35^ We agree that this is a useful approach because it reduces the complex problem of sub-group analysis into a single measure (reducing over-fitting), and because, by definition, it targets variables which most strongly predict the risk of target events. This latter aspect is important as it helps inform evidence synthesis models applying trial results to a target population with a higher target event rate. For similar reasons we propose that trials should also report evidence of treatment effect modification by comorbidity or degree of frailty; this would both reduce the risk of overfitting by reducing comorbidity to a single variable that predicts rates of competing events. To inform judgements about net benefits, this same information should be provided for adverse events. In addition, more research is required to establish whether specific comorbidities may attenuate or strengthen treatment efficacy, as if these effects were in the opposite direction for different comorbidities, then a cumulative count of comorbidities may obscure this effect.

## Conclusion

We found no evidence that treatment efficacy differed by comorbidity within the levels of comorbidity observed within clinical trial populations. This finding held whether comorbidity was measured using a simple condition count or by the presence or absence of six common conditions. The analysis of these trials may be used to inform subsequent evidence syntheses, analysis and reporting of individual trials, meta-analyses, and health economic models. We provide model outputs in the form of prior distributions to support such analyses.

## Data Availability

Aggregated data and code required to run these models, along with full model descriptions, are available at https://github.com/ChronicDiseaseEpi/csdr_effect_estimates

## Declarations

## Acknowledgements

This study, carried out under YODA Project # 2017-1746, used data obtained from the Yale University Open Data Access Project, which has an agreement with JANSSEN RESEARCH & DEVELOPMENT, L.L.C.. The interpretation and reporting of research using this data are solely the responsibility of the authors and does not necessarily represent the official views of the Yale University Open Data Access Project or JANSSEN RESEARCH & DEVELOPMENT, L.L.C.. This study was also carried out under ClinicalStudyDataRequest.com project number 1732, used data from the ClinicalStudyDataRequest.com repository, who provided data from Boehringer-Ingelheim, GSK, Lilly, Roche, Takeda, and Sanofi. The interpretation and reporting of research using these data are solely the responsibility of the authors and does not necessarily represent the official views of ClinicalStudyDataRequest.com or Boehringer-Ingelheim, GSK, Lilly, Roche, Takeda or Sanofi.

## Authors’ contributions

DM, PH, AS, BG, SD and NW conceived the study. DM acquired the data from trials and SAIL. PH, DM, EB and LH processed the data. DM and PH conducted the statistical analysis and interpretation of the data. NW and SD advised on the statistical analysis. PH wrote the first draft with support from DM. EB, AS, LH, SW, BG, FSM, DK, SD, NW and DM reviewed the manuscript and made critical changes for intellectual content. All authors read and approved the final manuscript.

## Ethical approval and consent to participate

This project had approval from the University of Glasgow, College of Medicine, Veterinary and Life Sciences ethics committee (200160070).

